# The impact of long-term non-pharmaceutical interventions on COVID-19 epidemic dynamics and control

**DOI:** 10.1101/2020.05.03.20089078

**Authors:** Marissa L. Childs, Morgan P. Kain, Devin Kirk, Mallory Harris, Lisa Couper, Nicole Nova, Isabel Delwel, Jacob Ritchie, Erin A. Mordecai

**Affiliations:** Emmett Interdisciplinary Program in Environment and Resources, Stanford University, Stanford, CA, 94305, USA; Department of Biology, Stanford University, Stanford, CA, 94305, USA; Natural Capital Project, Woods Institute for the Environment, Stanford University, Stanford, CA 94305, USA; Department of Zoology, University of British Columbia, Vancouver, BC V6T 1Z4, Canada; Department of Computer Science, Stanford University, Stanford, CA, 94305, USA

## Abstract

Non-pharmaceutical interventions to combat COVID-19 transmission have worked to slow the spread of the epidemic but can have high socio-economic costs. It is critical we understand the efficacy of non-pharmaceutical interventions to choose a safe exit strategy. Many current models are not suitable for assessing exit strategies because they do not account for epidemic resurgence when social distancing ends prematurely (e.g., statistical curve fits) nor permit scenario exploration in specific locations.

We developed an SEIR-type mechanistic epidemiological model of COVID-19 dynamics to explore temporally variable non-pharmaceutical interventions. We provide an interactive tool and code to estimate the transmission parameter, *β*, and the effective reproduction number, 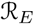. We fit the model to Santa Clara County, California, where an early epidemic start date and early shelter-in-place orders could provide a model for other regions.

As of April 22, 2020, we estimate an 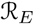 of 0.982 (95% CI: 0.849 - 1.107) in Santa Clara County. After June 1 (the end-date for Santa Clara County shelter-in-place as of April 27), we estimate a shift to partial social distancing, combined with rigorous testing and isolation of symptomatic individuals, is a viable alternative to indefinitely maintaining shelter-in-place. We also estimate that if Santa Clara County had waited one week longer before issuing shelter-in-place orders, 95 additional people would have died by April 22 (95% CI: 7 - 283).

Given early life-saving shelter-in-place orders in Santa Clara County, longer-term moderate social distancing and testing and isolation of symptomatic individuals have the potential to contain the size and toll of the COVID-19 pandemic in Santa Clara County, and may be effective in other locations.

## Introduction

COVID-19 is rapidly expanding across the globe and has the potential to overwhelm healthcare systems, killing hundreds of thousands to millions of people worldwide in the process^1^. Without an effective vaccine or specific drug therapy, non-pharmaceutical interventions such as physical distancing, diagnostic and serological testing, and contact-tracing are the best available tools to slow the spread of the pandemic and to mitigate its health toll. Governments and other decision-makers have used models to predict the spread of COVID-19 and show the benefits of social distancing for “flattening the curve,” i.e., slowing the epidemic—reducing and delaying the peak—to prevent medical systems from becoming overwhelmed.

Many decision-makers internationally, nationally, and locally, have used models that are statistical curve-fits, such as the IHME model^2^, to the observed numbers of COVID-19 cases, hospitalizations, or deaths, without capturing the underlying epidemiological dynamics of transmission. While statistical models can be successful at describing near-term epidemic trajectories, they may fail to capture the high degree of uncertainty in the long-term epidemic process, and therefore should not be used to project far into the future^3^. More worryingly, these models cannot anticipate impacts of major shifts in policy, such as ending shelter-in-place orders and reopening businesses. Thus, policy informed by statistical curve-fitting models may fail to anticipate the potential for a resurgence of COVID-19 epidemics, and therefore will not be able to adequately inform exit strategies from shelter-in-place and other social distancing interventions.

Epidemiological models that directly model the transmission process almost universally predict that lifting interventions too soon will result in a devastating resurgence in the epidemic^1^, a phenomenon supported by historical evidence, including data from the 1918 flu pandemic^4^. Balancing the economic and social costs of shelter-in-place orders with those of resurgence events, all of which are overwhelmingly borne by the most vulnerable, make identifying safe and effective exit strategies an urgent priority. However, many currently available epidemiological models are not set up for other scientists or policymakers to conveniently explore a variety of exit strategies for specific locations to which the model is also fit.

We developed an epidemiological compartment model of COVID-19 dynamics that uses a time-varying transmission parameter, *β*, to investigate the impact of non-pharmaceutical interventions on epidemic dynamics and control. The model incorporates transmission from both asymptomatic and presymptomatic infectious people. By fitting the model to local epidemic dynamics (using daily reported COVID-19 deaths), we can estimate key epidemiological metrics and evaluate the effectiveness of different long-term intervention strategies. Specifically, we explore three classes of strategies: 1) long-term shelter-in-place orders, which we consider the most drastic approach; 2) widespread testing and isolation of symptomatic people paired with less intensive social distancing in the general population; 3) an adaptive triggering approach that ramps up or turns down levels of social distancing when hospitalizations reach critical thresholds. We are particularly interested in identifying intervention strategies that do not require long-term sheltering-in-place while still maintaining epidemic control until a vaccine becomes widely available.

The goals of the model are to capture the transmission process accurately enough to understand qualitative impacts of intervention strategies without requiring extensive data on contact patterns, demography, movement, and other population features. We therefore model a homogeneous population with population-average parameters that reflect the demography of the population of interest. As a case study, we focus on Santa Clara County, California, where the first COVID-19 death in the U.S. was retroactively reported from February 6, 2020, and where the first-in-the-nation shelter-in-place order took effect early in the epidemic, on March 17, 2020. We estimate transmission rate for Santa Clara County under pre-intervention and shelter-in-place conditions, calculate reproduction numbers before and during interventions, explore the impact of long-term intervention strategies, and investigate counterfactuals to understand the impact of early intervention decisions. This case study illustrates how the model could be tailored to other locations to understand the impact of long-term interventions in COVID-19 epidemic dynamics.

## Methods

### Model Structure

We developed a compartmental model using an SEIR (Susceptible, Exposed, Infectious, Recovered) framework. We divided the population into states with respect to COVID-19: susceptible (S); exposed but not yet infectious (E); infectious and presymptomatic (I_P_), asymptomatic (I_A_), mildly symptomatic (I_M_), or severely symptomatic (I_S_); hospitalized cases that will recover (H_R_) or die (H_D_); recovered and immune (R); and dead (D), as shown in Equation sets S1 - S2; Figure S1. Parameters are defined in Tables 1 and 2.

By including asymptomatic and presymptomatic individuals, we are able to track “silent spreaders” of the disease, both of which have been shown to contribute to COVID-19 transmission^5^. Tracking hospitalizations and deaths allows us to compare our simulations to data sources that should be more reliable than confirmed cases, particularly in the absence of widespread rapid testing and case detection. Mildly symptomatic cases are defined as those people that show symptoms but do not require hospitalization, while we assume that all severely symptomatic cases will eventually require hospitalization. We also assume that no onward transmission occurs from hospitalized individuals.

The transmission parameter, *β*, describes the average per capita rate of contact between susceptible and infectious people multiplied by the per-contact transmission probability; we allow this parameter to vary over time to represent different social distancing strategies (i.e., stronger social distancing decreases *β* by decreasing the per capita rate of infectious contacts).

### Fitting the Model

We estimated both *β*_0_, which describes the initial value of *β* in the absence of any interventions, and *σ*, which describes the proportional reduction in *β*_0_ under shelter-in-place, where *β* = *β*_0_ · *σ*. To estimate *β*_0_ and *σ*, we assumed point estimates for some parameters (Table 1) and drew 200 sobol sequences across a range of plausible values for others (Table 2) to form 200 plausible parameter sets.

**Table 1:** Parameter point estimates.

| *Parameter* | *Value* | *Description* | *Citation* |
| --- | --- | --- | --- |
| $C_P, C_M, C_S$ | 1 | Relative infectiousness of presymptomatic, mild symptomatic, and severe symptomatic | Assumed |
| $\gamma$ | 3.5 days | Preinfectious period | 6,7 |
| $\lambda_P$ | 1.5 days | Presymptomatic duration | 8 |
| $\lambda_A$ | 7 days | Infectious period for asymptomatic infections | 9 |
| $\lambda_S$ | 5.5 days | Time from symptom onset to hospitalizations (severe cases) | 10,11 |
| $\lambda_M$ | 5.5 days | Time from symptom onset to recovery (mild cases) | 9 |
| $\rho_R$ | 13.3 days | Time from hospitalization to recovery | 12 |
| $\rho_D$ | 15 days | Time from hospitalization to death | 13 |
| N | 1.398x10<sup>6</sup> | Population | 14 |

**Table 2:**
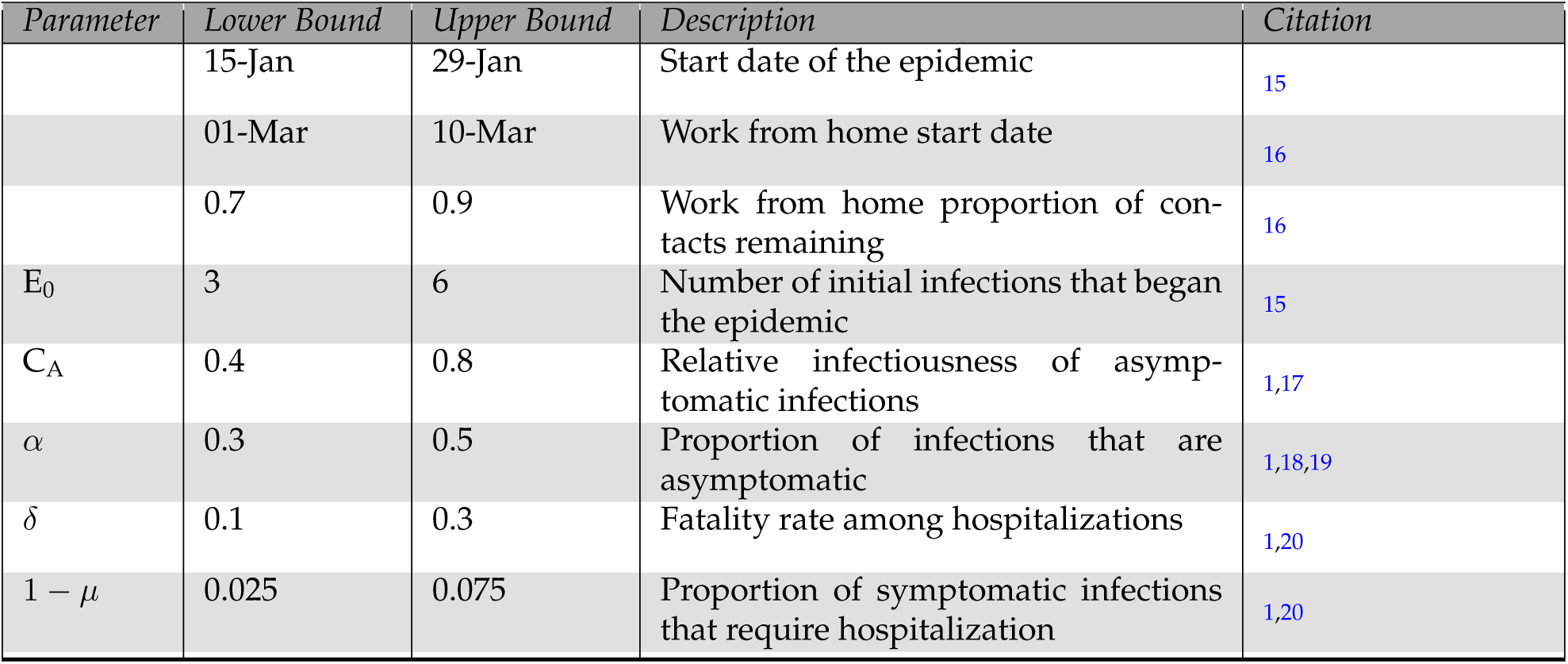
Parameter range estimates, some of which are specific to Santa Clara County, California.

Using the pomp package^21^ (function mif2) in the R programming language^22^, we fit both parameters to daily deaths for each of the 200 parameter sets using six particle filtering runs with variation in starting values; each run used 100 iterations and 3000 particles.

We use COVID-19 death data from The New York Times, based on reports from state and local health agencies^23^. Daily deaths are calculated from differences in cumulative death reports. Using these data, which are available for all counties in the US, our model can be used to fit *β*_0_ and *σ* in any county. Location-specific variation in these parameters results from differences in social structures, population immunity, population density, and other factors that determine the number of potentially infectious contacts and the per-contact transmission probability. For a given location, our model assumes that the population is homogeneous with a single average value for each parameter.

We calculated 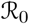 as estimated *β*_0_ times the duration of an average infection (as defined by our model structure) for each of the 200 parameter sets (using all six estimates from the mif2 iterations). We estimated 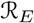 on April 22 using the estimated *β*_0_*, σ*, and the median proportion of the population remaining susceptible across the 300 simulated epidemics.

### Simulating epidemics under interventions

Our modeling framework allows for different types, intensities, and durations of interventions, and thereby illustrates how these interventions impact dynamics and the resulting number of COVID-19 cases and fatalities through time. We consider three possible interventions that can be implemented at different times during the simulation:

1. **Social distancing for a set duration** applied as a scaling of the transmission rate for all individuals
2. **Isolation of symptomatic individuals** applied as a scaling of the transmission rate for only symptomatic individuals I_S_ and I_M_; we assume isolation paired with partially relaxed social distancing
3. **Adaptive triggering** applied as a tightening or relaxing of social distancing, triggered by hospitalizations crossing a defined threshold

Other scenarios that can be modeled as a time-varying reduction in *β*_0_ can be explored using the code available on GitHub^24^.

To visualize the dynamics of a single intervention scenario, we simulate 300 epidemics from the single best fit across the 200 parameter sets as defined by negative log likelihood. To quantify the effectiveness of each intervention scenario, we estimate summary statistics from the simulated epidemics, such as total deaths, for a range of parameters for each intervention (e.g., the effectiveness of infected isolation). For each scenario, we simulate 300 epidemics across each of the 200 parameter sets, and calculate the 95% confidence interval (CI) for the summary statistic across all simulated epidemics. Here we define a 95% CI as an interval that captures the central 95% range of outcomes seen across all parameter sets and stochastic simulations. These are simultaneously wide because of large numbers of stochastic simulations, but narrow because we ignore uncertainty in all parameters listed in Table 1, and thus should be interpreted with caution.

## Results

### Local epidemic dynamics and control: Santa Clara County, California

We fit the model to Santa Clara County, California where work-from-home, social distancing, and shelter-in-place orders occurred early in the epidemic. We estimated that in the absence of controls, 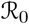 was 2.88 (95% CI: 2·47 - 3·45) in Santa Clara County, and that under our estimated efficacy of current shelter-in-place orders, 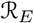 in Santa Clara as of April 22 is 0·98 (95% CI: 0·85 - 1·11) (Figure 1). We estimated 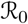 and 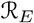 over time by holding out recent data to understand how our ability to estimate 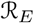 evolved as the epidemic unfolded (Figure S2). From stochastic simulations with the fitted parameter sets, we further estimated the percent of Santa Clara County population that would have been in the recovered class on April 22 (Figure S3).

**Figure 1:**
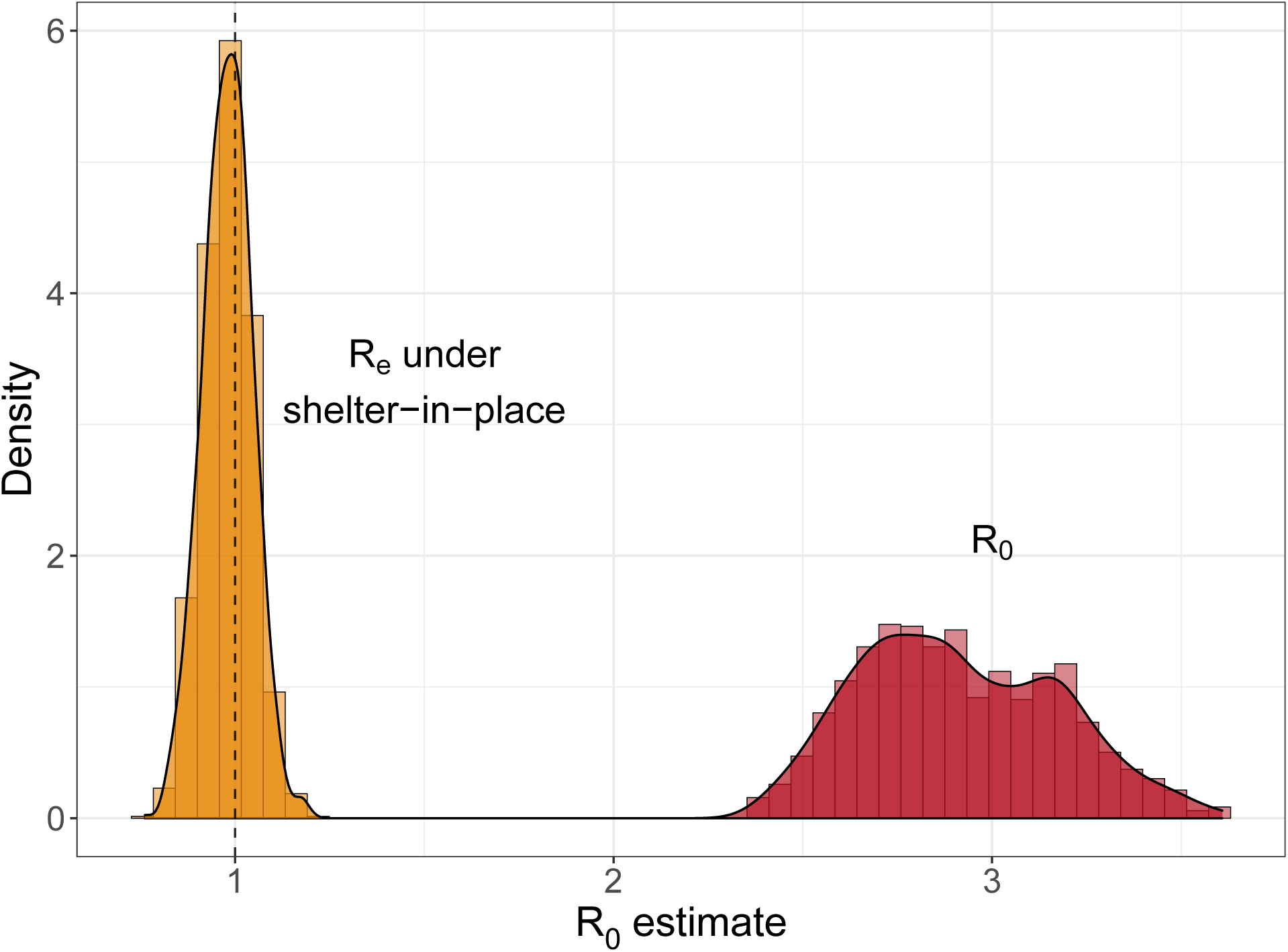
Distribution of 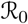 (red) and 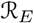 (gold) estimates in Santa Clara County, California as of April 22, 2020.

If shelter in place is simply lifted on June 1, 2020, we estimate that a second peak is inevitable in the absence of any non-pharmaceutical interventions, as illustrated here for one parameter set (Figure 2). Across all 200 parameter sets and stochastic epidemic simulations, we estimate a median of 5,478 deaths (95% CI: 1,767 - 11,632) and a peak number of concurrent infections of 171,667 (95% CI: 124,307 - 211,640) occurring on August 12 (95% CI: July 23 - September 9).

**Figure 2:**
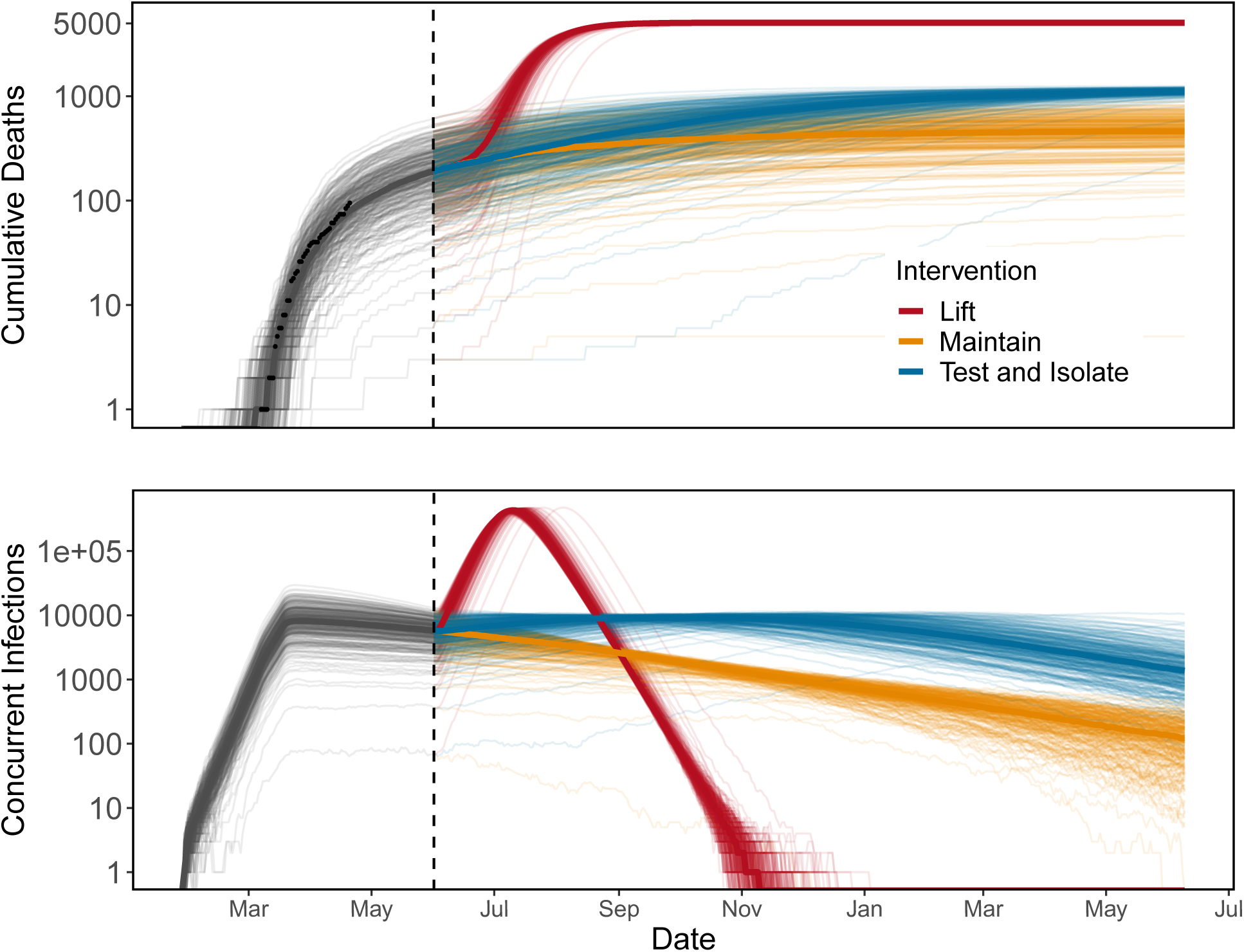
Maintaining shelter-in-place (gold) or test-and-isolate (blue) strategies over long periods are necessary to prevent a major epidemic resurgence (red) following the end of the initial shelter-in-place order on June 1 (dashed vertical line) in Santa Clara County. Lines show stochastic simulations of cumulative deaths (top, black points: observed data) and concurrent infections (bottom) for a single parameter set. Dates range from February 2020 (left) to July 2021 (right).

Maintaining shelter-in-place until June 1, followed by less stringent social distancing (50% of baseline contacts), combined with strong symptomatic case isolation (removing an additional 80% and 70% of invective contacts from severe and mild infections respectively), allows for higher background contact rates (e.g., more businesses reopening). For the parameter set shown, this scenario leads to an increase in mortality compared to maintaining shelter-in-place (Figure 2). Across a range of efficiencies of symptomatic case isolation in Santa Clara County, we find an overlap in CIs for deaths at all parameter sets but higher medians at the weakest levels of social distancing in the general population (Figure 3). For reference, the median number of estimated deaths under maintained shelter-in-place is shown by the horizontal black line, with 80% and 95% CI in dashed and dotted lines, respectively. These confidence intervals span a wide range because our estimated values range from 0·85 - 1·18, which leads to some epidemics growing and some declining through time.

**Figure 3:**
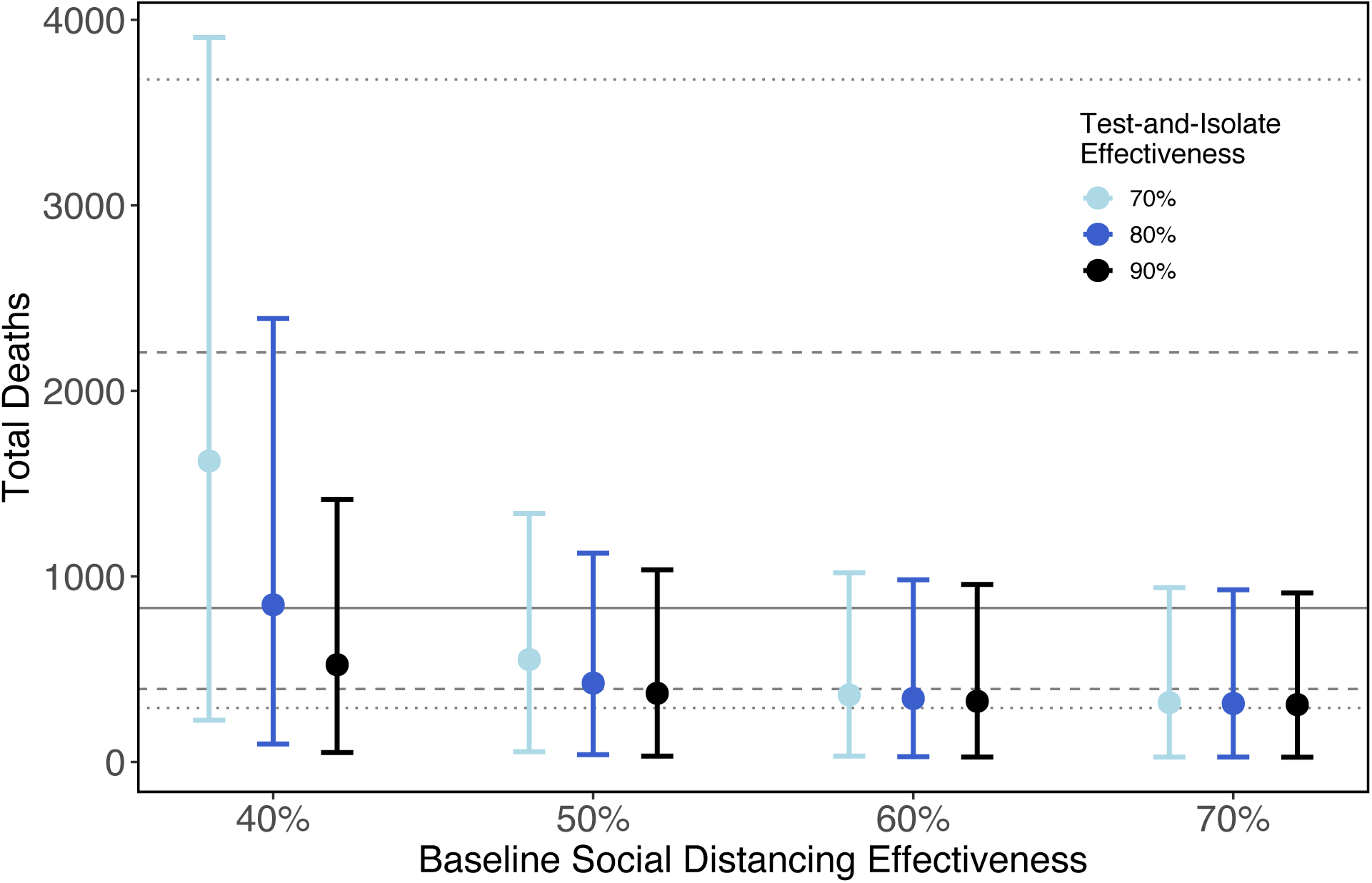
Similar total COVID-19 deaths are expected under various test-and-isolate strategies which include the effectiveness of symptomatic isolation (point colors), and social distancing effectiveness in the general population (x-axis). Due to both parameter uncertainty (here 100 randomly chosen from all 200 for computational reasons) and stochastic simulations, 95% CI are wide (point: median, error bars: 95% CI). Here, CI summarize cumulative deaths through June 2021. Epidemic toll begins to diverge when social distancing in the general public is weak (40% effectiveness) and symptomatic isolation is also weak (70% effectiveness). Lines represent the median (solid), 80% (dashed), and 95% (dotted) confidence intervals for maintaining current shelter-in-place orders indefinitely.

If widespread testing is not available before the end of shelter-in-place, a hypothetical alternative strategy is adaptive triggering, in which social distancing orders are intensified and relaxed as hospitalizations exceed and fall below critical thresholds. However, because the estimated 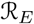 for Santa Clara County is approximately one (and CI spans one), a strategy that periodically reduces the strength of social distancing may lead to an overall increase in cases that is not reversed when the current shelter-in-place is reinstated. In the advent of more stringent shelter-in-place (e.g., reducing infectious contacts to 0·20 of baseline, which is realistic in other settings), an adaptive triggering strategy that alternates between a social distancing strength of 0·20 and 0·50 could be effective in keeping hospitalizations low (Figure S4). This method keeps the epidemic within the capacity of the healthcare system, but results in prolonged cycles of epidemic resurgence and control that continue until herd immunity is reached through recovery of infected individuals or vaccination.

## Counterfactuals

Santa Clara County’s early shelter-in-place order (enacted on March 17, 2020) helped to keep the death toll low; we estimate that waiting even one additional week would have led to an additional 95 deaths (95% CI: 7 - 283) by April 22, 2020. (Figure 4, orange trajectories and histogram). Alternatively, the implementation of test-and-isolate starting on March 17, 2020 in addition to the shelter-in-place (assuming an additional proportional reduction in contacts for mildly symptomatic and severely symptomatic infections by 0-3 and 0-2 respectively), would have helped to save an additional 24 lives (95% CI: 81 - [-1] (one extra death), green trajectories and histogram) (Figure 4).

**Figure 4:**
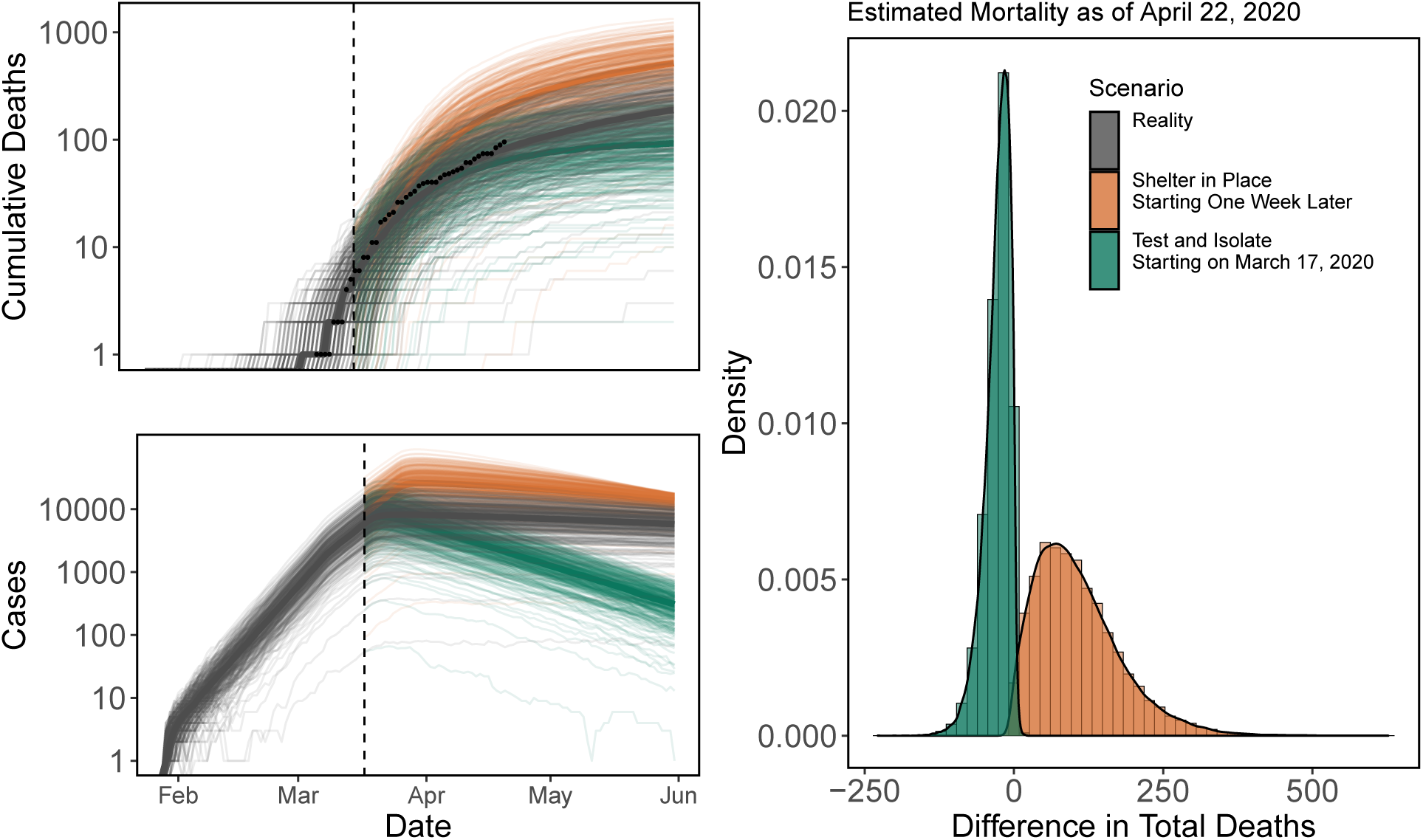
Early shelter-in-place in Santa Clara County saved lives, but early testing and isolation of symptomatic infections could have saved more. Cumulative deaths observed (top left panel; points) and daily cases (bottom left panel) under stochastic simulations using a single parameter set (lines; medians are in darker lines) under reality (gray), delaying shelter-in-place by one week (gold), or starting test-and-isolate on March 17, 2020 (green). We assume counterfactuals diverge on March 17, 2020 (vertical dashed line), the beginning of Santa Clara County shelter-in-place. Histograms (right) show the differences in the number of deaths for each stochastic model realization under the counterfactual scenarios compared to reality for all parameter sets.

## Discussion

Long-term interventions will be necessary to control the COVID-19 pandemic until more effective therapeutic drugs and vaccines are widely available: possibly 12-18 months from now, but potentially by the winter of 2020-2021. We found that social distancing orders such as work-from-home and shelter-in-place are effective at flattening the curve. However, lifting such measures even after periods of three to ten months or longer—depending on the strength of the intervention and the local transmission setting—risks allowing a major resurgence in the epidemic, undoing hard-won gains from social distancing measures. As an alternative to blanket shelter-in-place orders over long periods of time, we explored the efficacy of test-and-isolate and adaptive triggering methods for epidemic control. We found that test-and-isolate measures paired with lighter social distancing, especially when combined with early shelter-in-place orders, can be effective at keeping the epidemic under control while presumably alleviating some of the social and economic costs of shelter-in-place. Given the social and economic challenges of maintaining shelter-in-place for months at a time, test-and-isolate interventions are a potential alternative until better therapeutics become widely available. Improved testing coverage would also have the added benefit of: 1) improving implementation of contact tracing to identify and quarantine contacts before they potentially become asymptomatic and presymptomatic spreaders, and 2) helping to fit models and other public health surveillance tools to COVID-19 cases, rather than deaths.

Recent evidence suggests that a large proportion of infected people may be asymptomatic or presymptomatic^25,26^, and that a larger proportion of the population than previously understood may have already been infected (Stanford seroprevalence study:^27^; Harvard seroprevalence study:^28^). Our model currently estimates that 1.18% (95% CI: 0-01% - 4-65%) of the Santa Clara County population has already recovered from infection, as of April 22, 2020. A better general understanding of the total magnitude of the epidemic size, based on improved diagnostic and serological testing, will help to tailor estimates of epidemic trajectories under different intervention scenarios, and to improve estimates of epidemiological parameters like 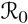.

The model we present is deliberately simplified in several respects so that we are able to use it in different settings, explore a range of intervention scenarios, and to fit using death data. For example, the model ignores heterogeneity in susceptibility, contact rates, and disease outcomes arising from population demographic structure, co-morbidities, mobility, and other factors. Additionally, we did not take into account hospital capacity, meaning that lifting interventions could potentially lead to more deaths than predicted here if capacity is overwhelmed and mortality rates increase. However, with this simple model and accompanying open-access code^24^ and interactive tool (`covid-measures.stanford.edu`) as a baseline for exploring qualitative long-term intervention scenarios, we expect that researchers and public health experts could adapt the model based on further data availability or locally-specific goals, either by adding state variables or adjusting intervention scenarios and parameter values.

Despite its simplicity, the model captures the early dynamics of COVID-19 in Santa Clara County well (Figure 2), and provides estimates of 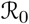 that broadly match other estimates in the literature^29^. The model clearly shows that early action in California, including work-from-home and shelter-in-place orders, saved lives. The qualitative effect of early social distancing on epidemic dynamics is robust to a wide range of parameter uncertainty; we estimated that waiting even one additional week to implement shelter-in-place would have led to an additional 7-283 deaths in the county by April 22, 2020. With the benefit of early action, some increase in social contact in the general public may be possible by June 1, given that the capacity for testing and isolation of symptomatic people continues to increase. Though we find that adaptive triggering is unlikely to work based on current conditions in Santa Clara County, it may be a viable option in locations such as Italy or India where legally-enforced lockdown has led to more stringent reductions in social contacts^30^. Expansion of diagnostic testing capacity is a top priority for long-term COVID-19 mitigation efforts because of its multifaceted benefits for concentrating social distancing efforts on those most at risk of transmitting COVID-19, for determining the true size and trajectory of epidemic dynamics, and for providing more certainty to individuals experiencing COVID-19 symptoms.

During an unfolding pandemic, modeling is an essential tool for tactical decision-making, strategic planning, and communication of qualitative scenarios to the public. The number of COVID-19 models has grown apace with the pandemic itself, and many of these models have overlapping goals and approaches; organizations such as the MIDAS Network (Models of Infectious Disease Agent Study) provide an important service in coordinating data-gathering and modeling efforts and in providing publicly available resources for the modeling community^29^. The rapid adoption of open-data policies from across the spectrum of academic, government, business, and media organizations has been a major boon to research and pandemic control efforts. At the same time, keeping up with the growing COVID-19 modeling literature is nearly impossible, and the differences among models remain confusing to the public amidst a fragmented pandemic response across US states, counties, and the federal government. While statistical curve-fitting models may be valuable for modeling short-term trajectories in cases, hospitalizations, deaths, and healthcare capacities^2,3^, epidemiological models that capture the underlying transmission dynamics are critical for evaluating the impact of major changes in policy over the long term. Epidemiological compartment models vary widely in how they subdivide populations and in the assumptions that govern movement among compartments. Individual-based, network, and meta-population models expand on compartmental models by capturing elements of individual and population heterogeneity that influence epidemic dynamics, but require much more extensive data for parameterization e.g., see^1^. Along the modeling continuum from statistical curve-fitting to compartmental models to individual-based, network, and meta-population models, compartmental models such as the one presented here are most useful for exploring long-term impacts of intervention scenarios across different settings where highly detailed data are not available.

We aimed to provide an open-source modeling tool that is detailed enough to capture key elements of transmission, including asymptomatic and presymptomatic transmission and a time-varying transmission coefficient, while remaining simple enough to be parameterized using widely available information and data. As demonstrated here, this relatively simple model captures key epidemiological dynamics and parameters in Santa Clara County, California, and suggests important qualitative differences among intervention scenarios. In settings where more detailed modeling tools and data are not readily available, this modeling approach can provide some guidance about qualitative impacts of different scenarios, and can be easily tailored to fit local epidemic dynamics. Most importantly, this model suggests that early interventions have already saved lives, and that exit strategies from shelter-in-place orders should be made thoughtfully and based on rigorous epidemiological models.

## Data Availability

Data and code used to produce the results in this study are available on github.

https://github.com/nytimes/covid-19-data

## Data and Code Availability

Data used in this study are available at https://github.com/nytimes/covid-19-data. Code used to produce the results in this study are available at https://github.com/morgankain/COVID_interventions.

## Acknowledgements

Funding provided by: the National Science Foundation (DEB-1518681); the National Institute of General Medical Sciences (1R35GM133439-01); the Natural Capital Project; the Helman Scholarship; the Terman Award. Marissa Childs was supported by the Illich-Sadowsky Fellowship through the Stanford Interdisciplinary Graduate Fellowship program at Stanford University. Morgan Kain was supported by the Natural Capital Project. Nicole Nova was supported by the Stanford Data Science Scholarship. Jacob Ritchie was supported by the Terry Winograd Fellowship. Mallory Harris was supported by the Knight-Hennessy Scholarship.

## Author Contributions

MC helped to design and code the model, conduct model simulations, troubleshoot problems and update the model, write and revise the manuscript.

MPK helped to design and code the model, conduct model simulations, troubleshoot problems and update the model, write and revise the manuscript.

DK helped to design the model, search the literature for parameter values and databases for data, troubleshoot problems and update the model, write and revise the manuscript.

MH helped to design the model, search the literature for parameter values and databases for data, troubleshoot problems and update the model, write and revise the manuscript.

LC helped to design the model, search the literature for parameter values and databases for data, and revise the manuscript.

NN helped to design the model, search the literature for parameter values and databases for data, and revise the manuscript.

ID helped to design the model, search the literature for parameter values and databases for data, and revise the manuscript.

JR created the public-facing website associated with our work, and helped revise the manuscript.

EAM helped to design the model, provide conceptual framing, and write and revise the manuscript.

## Supplement: Equations and model implementation

We assume an underlying, unobserved process model of SARS-CoV-2 transmission described by Equation set S1 and shown in Figure S1. The compartments in the model are susceptible (*S*); exposed but not infectious (*E*); infectious and asymptomatic (*I_A_*), presymptomatic *(I_P_*), mildly symptomatic *(I_M_*), and severely symptomatic (*I_S_*); hospitalized cases that will recover (*H_R_*) or die *(H_D_*); and recovered (*R*). We use an Euler approximation of the continuous time process with a time step of 4 hours. Transitions between compartments are simulated as binomial (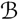) or multinomial (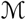) processes; Equation set S2 describes in detail the stochastic rates used to approximate the transition terms in Equation set S1. Parameters are defined in Tables 1 and 2. Finally, we assume that the observed deaths are a Poisson random variable with mean of total new deaths accumulated over the observation period (i.e. one day for this analysis).

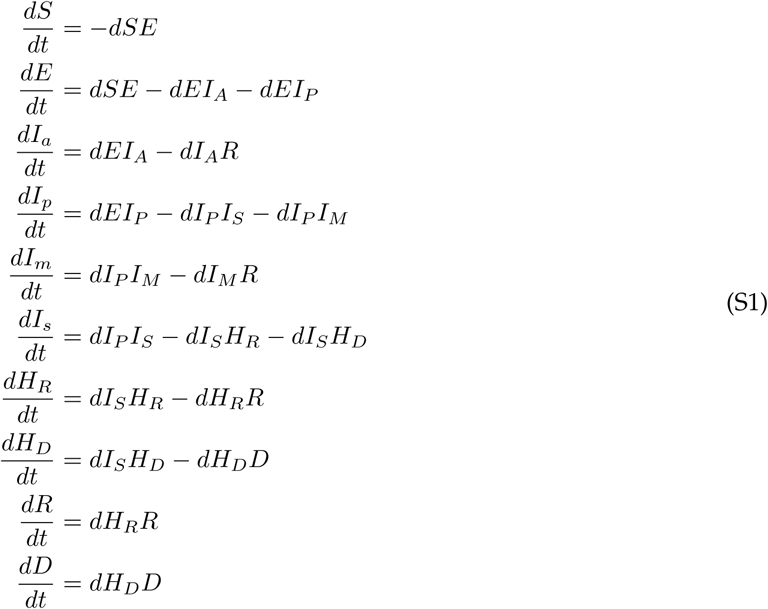

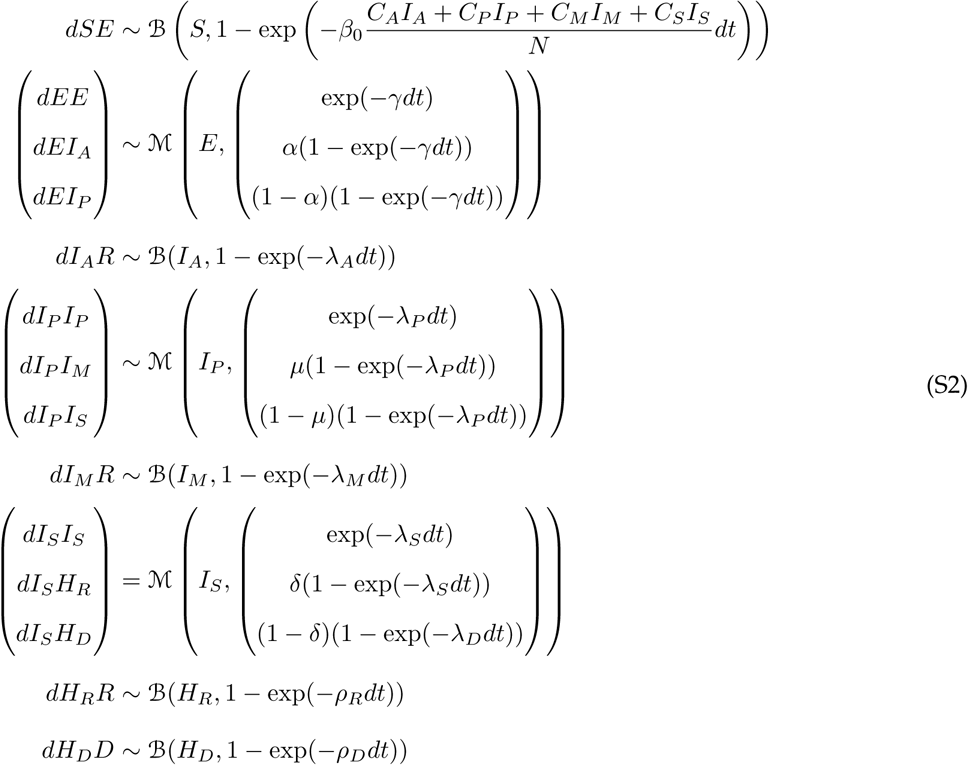

Supplement: Figures

**Figure S1:**
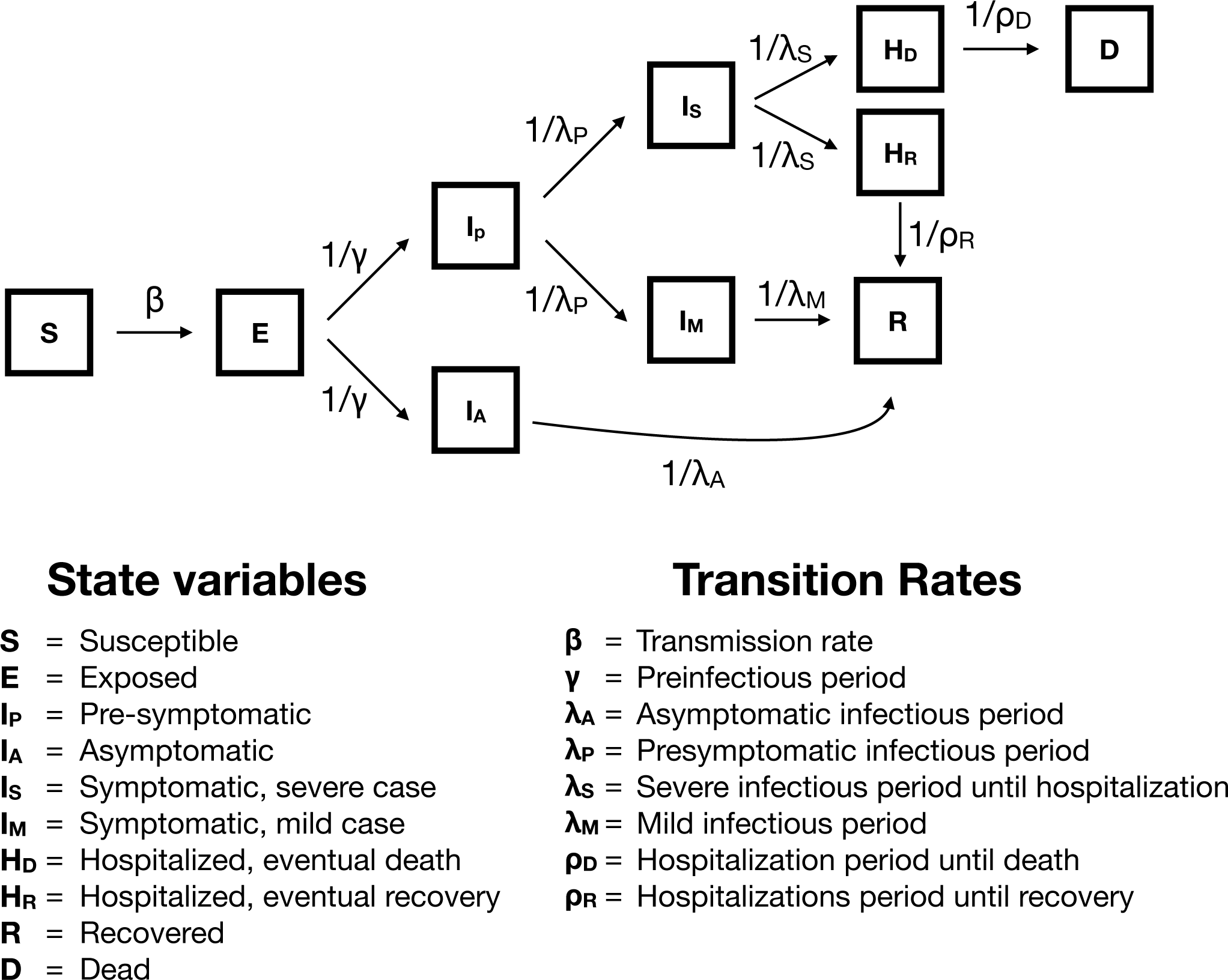
Epidemiological model box diagram

**Figure S2:**
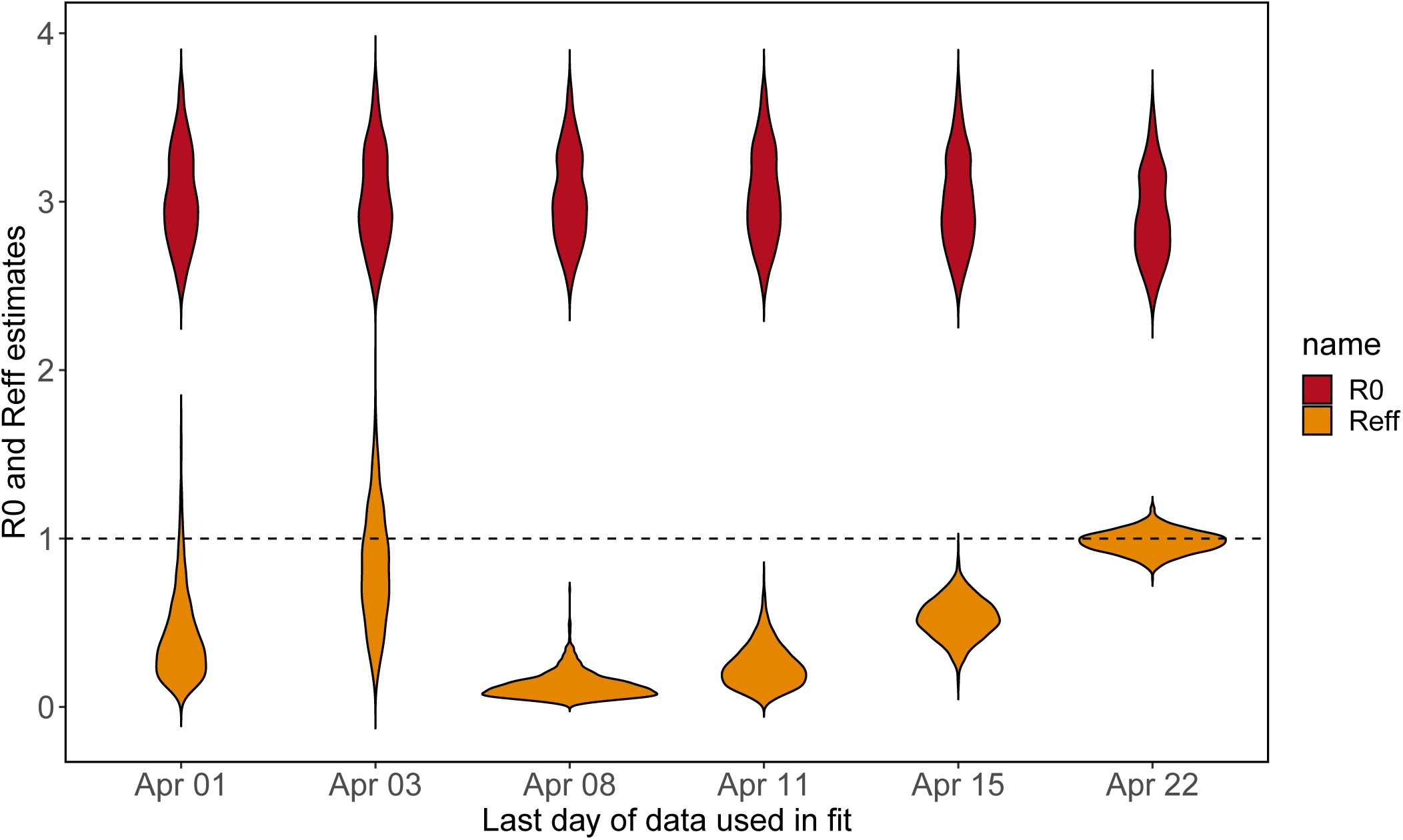
Estimated 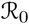 and 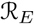 from fits to truncated time series. A date corresponding to a pair of violin plots shows the most recent data for which data was used to fit the model. The low and confident 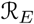 estimate on April 8 was due in part to 5 consecutive days, ending on April 8, with a total of 3 deaths.

**Figure S3:**
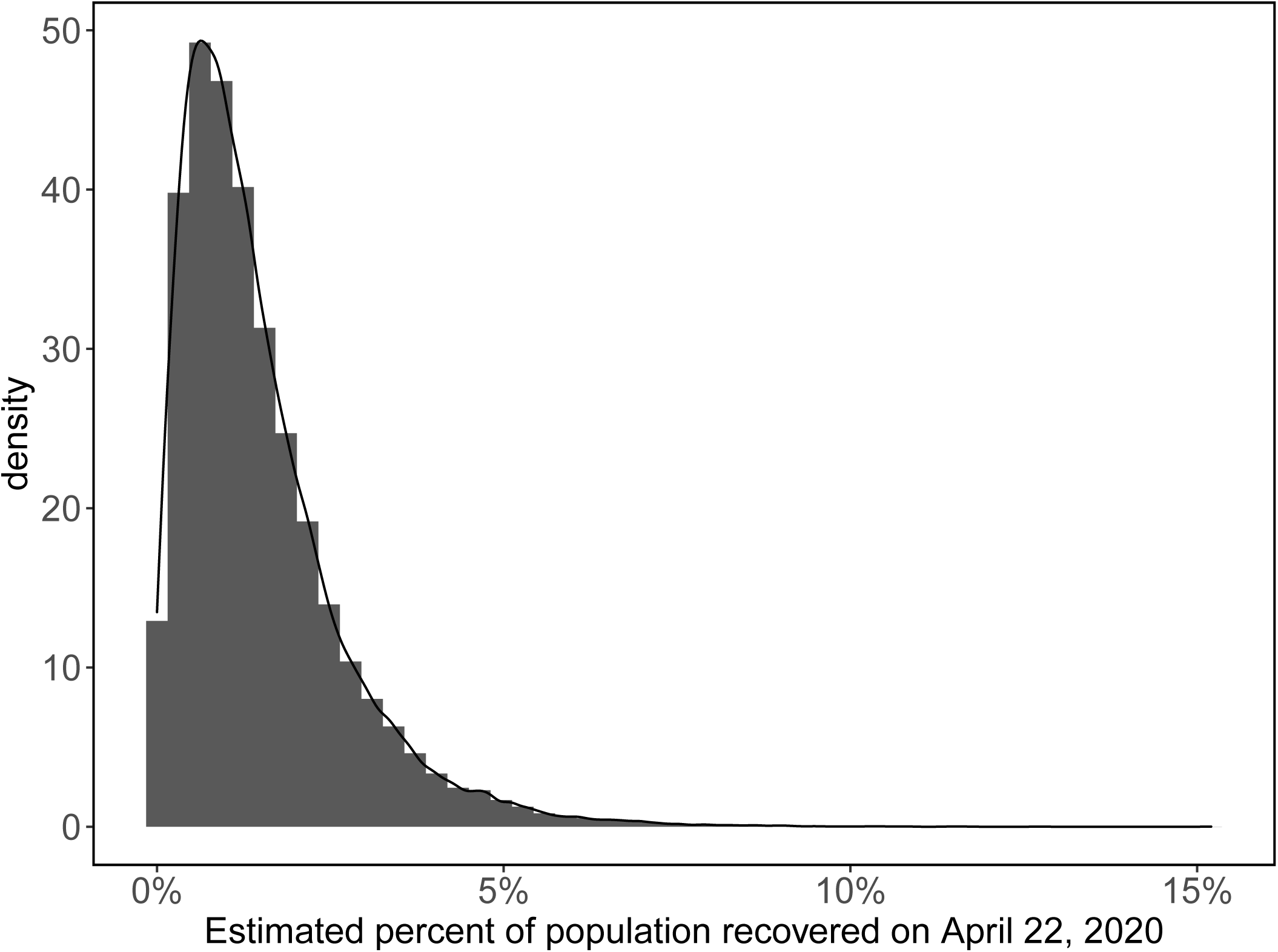
Distribution of estimated percent of Santa Clara County in the recovered class from 300 simulations of 200 parameter sets.

**Figure S4:**
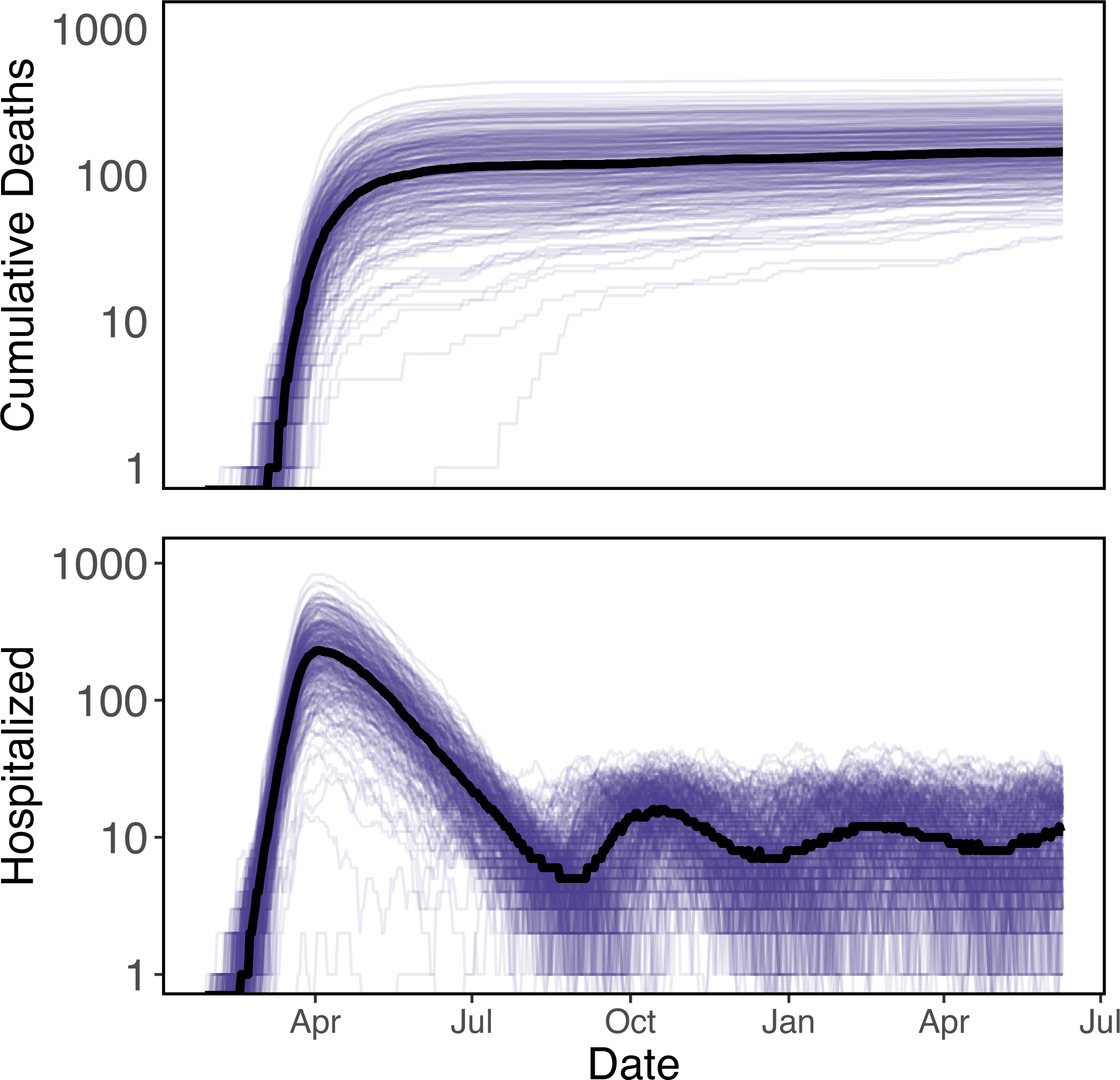
Adaptive triggering that alternates between a social distancing strength of 20% of background contacts and 50% of background contacts when the number of people hospitalized reaches 15 people or falls to 5 people, respectively. This strategy results in a moderately constant number hospitalized and leads to a slowly increasing cumulative death toll over time.

## References

[1] Ferguson N, Laydon D, Nedjati Gilani G, Imai N, Ainslie K, Baguelin M, et al. Report 9: Impact of non-pharmaceutical interventions (NPIs) to reduce COVID19 mortality and healthcare demand; 2020. Available from: http://spiral.imperial.ac.uk/handle/10044/1/77482.

[2] IHME COVID-19 Projections;. Available from: https://covid19.healthdata.org/ (accessed April 23, 2020).

[3] Woody S, Garcia Tec M, Dahan M, Gaither K, Lachmann M, Fox S, et al. Projections for first-wave COVID-19 deaths across the US using social-distancing measures derived from mobile phones. medRxiv. 2020;Available from: https://www.medrxiv.org/content/early/2020/04/26/2020.04.16.20068163.

[4] Hatchett RJ, Mecher CE, Lipsitch M. Public health interventions and epidemic intensity during the 1918 influenza pandemic. Proceedings of the National Academy of Sciences. 2007;104(18):7582–7587.

[5] Ferretti L, Wymant C, Kendall M, Zhao L, Nurtay A, Abeler-Döner L, et al. Quantifying SARS-CoV-2 transmission suggests epidemic control with digital contact tracing. Science. 2020; DOI:10.1126/science.abb6936.

[6] Lauer SA, Grantz KH, Bi Q, Jones FK, Zheng Q, Meredith HR, et al. The Incubation Period of Coronavirus Disease 2019 (COVID-19) From Publicly Reported Confirmed Cases: Estimation and Application. Annals of Internal Medicine. 2020; Available from: https://doi.org/10.7326/M20-0504.

[7] Zhang J, Litvinova M, Wang W, Wang Y, Deng X, Chen X, et al. Evolving epidemiology and transmission dynamics of coronavirus disease 2019 outside Hubei province, China: a descriptive and modelling study. The Lancet Infectious Diseases. 2020; Available from: http://www.sciencedirect.com/science/article/pii/S1473309920302309.

[8] Wei WE, Li Z, Chiew CJ, Yong SE, Toh MP, Lee VJ. Presymptomatic Transmission of SARS-CoV-2—Singapore, January 23-March 16, 2020. Morbidity and Mortality Weekly Report. 2020;69(14):411.

[9] Wolfel R, Corman VM, Guggemos W, Seilmaier M, Zange S, Müller MA, et al. Virological assessment of hospitalized patients with COVID-2019. Nature. 2020 Apr;p. 1–5.

[10] Sanche S, Lin YT, Chonggang Xu, Ethan Romero-Severson, Nick Hengartner, Ruian Ke. High Contagiousness and Rapid Spread of Severe Acute Respiratory Syndrome Coronavirus 2. Emerging Infectious Disease journal. 2020;26(7). Available from: https://www.nc.cdc.gov/eid/article/26/7/20-0282_article.

[11] Han Yn, Feng Zw, Sun Ln, Ren Xx, Wang H, Xue Ym, et al. A comparative-descriptive analysis of clinical characteristics in 2019-coronavirus-infected children and adults. Journal of Medical Virology;Available from: https://onlinelibrary.wiley.com/doi/abs/10.1002/jmv.25835.

[12] Tindale L, Coombe M, Stockdale J, Garlock E, Lau WYV. Transmission interval estimates suggest presymptomatic spread of COVID-19 | medRxiv; 2020. Available from: https://www.medrxiv.org/content/10.1101/2020.03.03.20029983v1.

[13] Gaythorpe K, Imai N, Cuomo-Dannenburg G, Baguelin M, Bhatia S, Boonyasiri A, et al. Report 8: Symptom progression of COVID-19. Imperial College London; 2020. Available from: http://spiral.imperial.ac.uk/handle/10044/1/77344.

[14] Bureau, US Census. Small Area Health Insurance Estimates (SAHIE) Program;. Library Catalog: www.census.gov Section: Government. Available from: https://www.census.gov/programs-surveys/sahie.html.

[15] Hadfield J, Bedford T, Neher R, Hodcroft E. Nextstrain: real-time tracking of pathogen evolution. Bioinformatics. 2018;.

[16] Descartes Labs. descarteslabs/DL-COVID-19. Descartes Labs; 2020. Original-date: 2020-03-25T18:11:21Z. Available from: https://github.com/descarteslabs/DL-COVID-19.

[17] Perkins A, Cavany SM, Moore SM, Oidtman RJ, Lerch A, Poterek M. Estimating unobserved SARS-CoV-2 infections in the United States. medRxiv. 2020;p. 2020.03.15.20036582. Available from: https://www.medrxiv.org/content/10.1101/2020.03.15.20036582v2.

[18] Lavezzo E, Franchin E, Ciavarella C, Cuomo-Dannenburg G, Barzon L, Vecchio CD, et al. Suppression of COVID-19 outbreak in the municipality of Vo, Italy. medRxiv. 2020 Apr;p. 2020.04.17.20053157. Publisher: Cold Spring Harbor Laboratory Press. Available from: https://www.medrxiv.org/content/10.1101/2020.04.17.20053157v1.

[19] Nishiura H, Linton NM, Akhmetzhanov AR. Serial interval of novel coronavirus (COVID-19) infections. International Journal of Infectious Diseases. 2020 Apr;93:284–286. Publisher: Elsevier. Available from: https://www.ijidonline.com/article/S1201-9712(20)30119-3/abstract.

[20] Verity R, Okell LC, Dorigatti I, Winskill P, Whittaker C, Imai N, et al. Estimates of the severity of coronavirus disease 2019: a model-based analysis. The Lancet Infectious Diseases. 2020;0(0). Available from: https://www.thelancet.com/journals/laninf/article/PIIS1473-3099(20)30243-7/abstract.

[21] King A, Ionides E, Breto C, Ellner S, Ferrari M, Kendall B, et al. pomp: Statistical Inference for Partially Observed Markov Processes; 2020. Available from: https://kingaa.github.io/pomp/.

[22] R Core Team. R: A Language and Environment for Statistical Computing. Vienna, Austria; 2017. Available from: https://www.R-project.org/.

[23] Coronavirus (Covid-19) Data in the United States. GitHub; 2020. https://github.com/nytimes/covid-19-data (accessed April 23, 2020).

[24] Kain MP, Childs ML. COVID-Interventions. GitHub; 2020. https://github.com/morgankain/COVID_interventions.

[25] Stock JH, Aspelund KM, Droste M, Walker CD. Estimates of the Undetected Rate among the SARS-CoV-2 Infected using Testing Data from Iceland. medRxiv. 2020;Available from: https://www.medrxiv.org/content/early/2020/04/11/2020.04.06.20055582.

[26] Song H, Xiao J, Qiu J, Yin J, Yang H, Shi R, et al. A considerable proportion of individuals with asymptomatic SARS-CoV-2 infection in Tibetan population. medRxiv. 2020;Available from: https://www.medrxiv.org/content/medrxiv/early/2020/03/30/2020.03.27.20043836.

[27] Bendavid E, Mulaney B, Sood N, Shah S, Ling E, Bromley-Dulfano R, et al. COVID-19 Antibody Seroprevalence in Santa Clara County, California. medRxiv. 2020;p. 2020.04.14.20062463. Available from: https://www.medrxiv.org/content/10.1101/2020.04.14.20062463v1.

[28] Larremore DB, Fosdick BK, Bubar KM, Zhang S, Kissler SM, Metcalf CJE, et al. Estimating SARS-CoV-2 seroprevalence and epidemiological parameters with uncertainty from serological surveys. medRxiv. 2020;Available from: https://www.medrxiv.org/content/medrxiv/early/2020/04/20/2020.04.15.20067066.

[29] MIDAS Online Portal for COVID-19 Modeling Research; 2020. https://midasnetwork.us/covid-19/ (accessed March 20, 2020).

[30] Lazzerini M, Putoto G. COVID-19 in Italy: momentous decisions and many uncertainties. The Lancet Global Health. 2020 May;8(5):e641–e642.

